# Lifting mobility restrictions and the induced short-term dynamics of COVID-19

**DOI:** 10.1101/2020.07.23.20161026

**Authors:** Mario Santana-Cibrian, Manuel A. Acuña-Zegarra, Jorge X. Velasco-Hernandez

## Abstract

SARS-CoV-2 has now infected 15 million people and produced more than six hundred thousand deaths around the world. Due to high transmission levels, many governments implemented social-distancing measures and confinement with different levels of required compliance to mitigate the COVID-19 epidemic. In several countries, these measures were effective, and it was possible to flatten the epidemic curve and control it. In others, this objective was not or has not been achieved. In far to many cities around the world rebounds of the epidemic are occurring or, in others, plateau-like states have appeared where high incidence rates remain constant for relatively long periods of time. Nonetheless, faced with the challenge of urgent social need to reactivate their economies, many countries have decided to lift mitigation measures at times of high incidence. In this paper, we use a mathematical model to characterize the impact of short duration transmission events within the confinement period previous but close to the epidemic peak. The model describes too, the possible consequences on the disease dynamics after mitigation measures are lifted. We use Mexico City as a case study. The results show that events of high mobility may produce either a later higher peak, a long plateau with relatively constant but high incidence or the same peak as in the original baseline epidemic curve, but with a post-peak interval of slower decay. Finally, we also show the importance of carefully timing the lifting of mitigation measures. If this occurs during a period of high incidence, then the disease transmission will rapidly increase, unless the effective contact rate keeps decreasing, which will be very difficult to achieve once the population is released.

## 1 Introduction

Latin America has recently become the world’s epicenter for SARS-CoV-2 activity. Lack of infrastructure, weak economic performance and social inequality can, in part, explain this situation. In many countries such as Mexico, a large percentage of the economically active population works in informal jobs that largely depend on going into the street and plazas of towns and cities. This explains the need to reopen the economy in these countries but, on the other hand, ineffectively justifies the lifting of social-distancing policies in the midst of the epidemic. However, not only the economically deprived countries of the American continent are suffering the blunt of the COVID-19 pandemic. The United States has been unable to bring under control the spread of the disease. As has been the case in many Latin American countries, the USA, in general, reopened its economy too fast and without a clear strategy in place. This has brought a worrisome rebound of the epidemic that threatens the very way of life of this rich and powerful country. It is within this context that we present our work. There is the need for strategic modeling, geared to local, specific situations, that can provide projections to understand and evaluate the consequences of lifting mitigation measures that were put into place earlier this year, to fight the so-called first epidemic wave at the local (county or city) levels. Mathematical modeling has been a valuable tool to understand the dynamics of COVID-19 around the world. Many models have employed variations of the SEIR model [1, 2, 3, 4, 5, 6]. In this work we particularly concentrate on the impact of what we call atypical mobility events, that are and were associated with population events where some level of disease superspreading occurred as previously observed in [7]. These short-duration, superspreading events have a non-negligible effect upon the shape and time evolution of the epidemic curve. In many places of the world, such as in the USA and, at present, in several regions of Mexico, we are seeing that the epidemic has entered a plateau, and when the plateau ends it gives place to a new epidemic growth phase. We exemplify our model applying it to the case of Mexico City. We analyze the interrelation between the date of maximum incidence and heightened incidence events taking place before the projected maximum. The manuscript is organized as follows. In Section 2, we formulate the mathematical model and give some general results arising from it; in Section 3, we describe the Mexico City data that is used to exemplify our model results and present a brief analysis of the impact of mitigation measures. In Section 4, we use our proposed model to generate scenarios that describe the impact of superspreading events associated with holidays, on the shape of the epidemic curve, and explore the probable consequences of lifting mitigation measures. Finally, the discussion and conclusions are presented in Section 5.

## 2 Mathematical Model Formulation

Our model is an extension of the mathematical model first introduced in [5]. We extend it as follows: i) we introduce a compartment for the *reported* infectious cases, ii) we introduce two time-dependent contact rates to approximate their reduction after the initial mitigation measures were implemented, and iii) we introduce a time-dependent confinement-compliance rate. These extensions allow us to provide a better account of the social-distancing effects and the role and impact of several short periods of increased transmission due to the lack of compliance of social-distancing guidelines during the confinement.

Our mathematical model explicitly takes into account two time intervals, before and after the date when mitigation measures were implemented, *T*, as illustrated in Figure 1. The model is of SEIR-type with three infectious classes: *I*_*a*_ asymptomatically, *I*_*s*_ symptomatically, and *I*_*r*_ reported (confirmed) infectious individuals. The dynamics for the first time interval are described by the following system of ordinary differential equations:

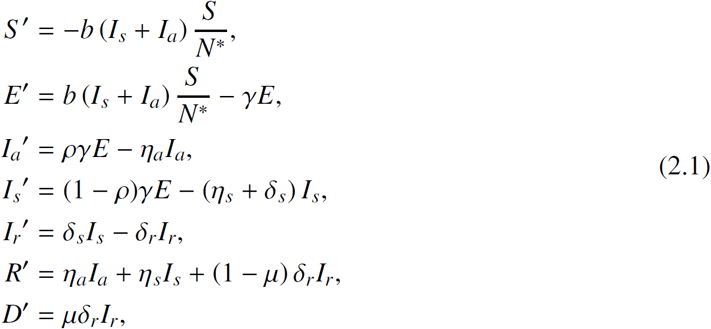

**Figure 1:**
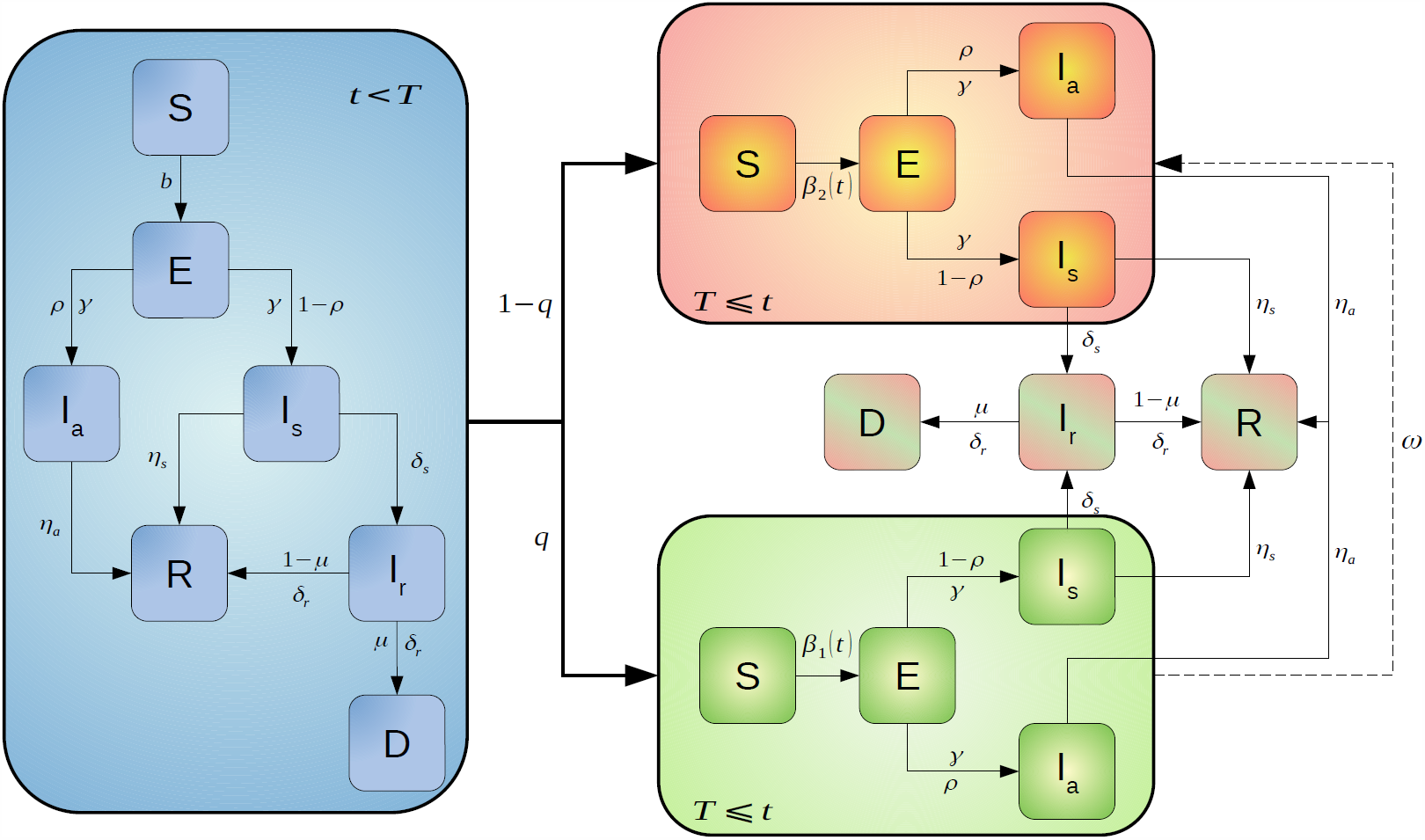
*S, E, I*_*a*_, *I*_*s*_, *I*_*r*_, *R, D* represent the populations of susceptible, exposed, asymptomatically infected, symptomatically infected, reported infected, recovered and dead individuals, respectively. Previous to the mitigation measures, the epidemic follows the dynamics represented by the blue diagram. Once the mitigation measures are implemented (on March 23 in the case of Mexico), the population splits into two: those who comply with the control measures (green box) and those who do not (orange box). The dashed line connecting the green and orange boxes represent the compliance-failure rate *ω*(*t*).

where *N*^∗^ = *S* + *E* + *I*_*a*_ + *I*_*s*_ + *R*. Note that the *I*_*r*_ compartment does not participate in transmission since, it is assumed, once confirmed, the cases are effectively isolated. *b* represents the effective contact rate, 1*/γ* the incubation period, *ρ* the proportion of asymptomatically infected individuals, 1*/η*_*a*_ and 1*/η*_*s*_ the periods from symptoms onset to recovery for symptomatically and asymptomatically infected individuals, *δ*_*s*_ the rate at which a symptomatically infected individuals becomes a reported (confirmed) case, 1*/δ*_*r*_ the time from confirmation to recovery or death and *µ* is the proportion of those reported cases that die. The basic reproductive number of the system 2.1 is given by:

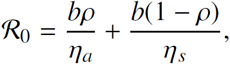

where the first term represents the number of new cases produced by asymptomatically infected individuals and the second term represents the number produced by symptomatically infected individuals. After mitigation measures are implemented (*T*), the population is split into two groups: one constituted by those individuals who comply with the measures that is referred to as the *confined group*, and another constituted by those individuals who do not, that is called the *unconfined group*, either because they disobey the social-distancing guidelines or because they belong to an strategic sector of the economy. The dynamics in the second time interval are given by:

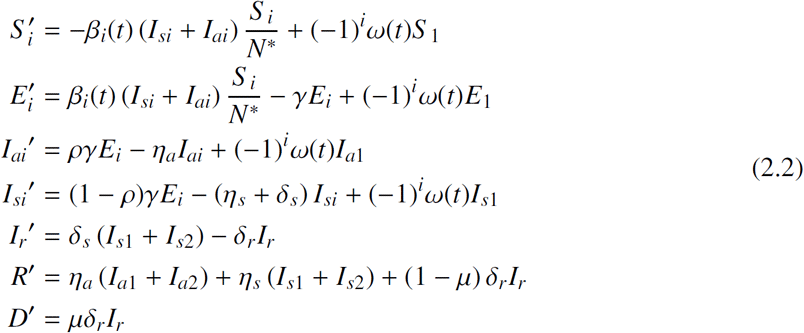

where 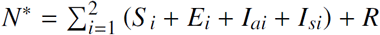. The index *i* = 1 gives the population that complies with the control measures, while *i* = 2 indicates those who do not. Note that the compartments *I*_*r*_, *R*, and *D* are common to both groups. Following [5], the effective contact rates are different for each group and are given by:

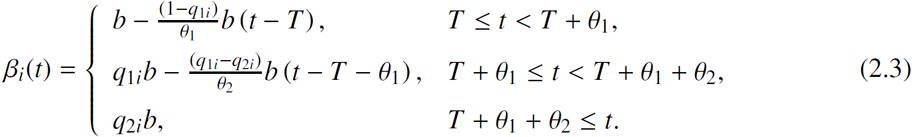

Equation (2.3) assumes decreasing contact rates where the parameters *q*_2*i*_ ≤*q*_1*i*_, *i* = 1, 2, represent a reduction in the baseline effective contact rate *b. T* is the time at which the effective contact rates start to act and *θ* _*j*_, *j* = 1, 2, are the duration of the contact reduction process. Finally, we define the compliance-failure rate *ω*(*t*) as a step function, that is,

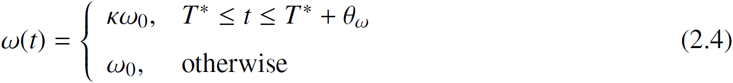

where *κ >* 1 is the increase in the compliance-failure rate relative to the baseline value *ω*_0_ (*κ* = 1), *T* ^∗^ is the time at which the perturbation starts and *θ*_*ω*_ is the duration of the perturbation. This model admits several periods during which *ω*(*t*) may act but here we illustrate only one period.

The inclusion of short-term superspreading events into equations 2.3 and 2.4 renders scenarios in which a plateau-like behavior appears. We show three scenarios: (I) long plateau with slight decrease and rebound; (II) shorter plateau with a later decreasing trend in the epidemic-curve; (III) long plateau with a later decreasing epidemic-curve. Each scenario is subdivided into three sub-cases defined as follows:

- Scenario X.a: Increase *κ* = 3 of the baseline value *ω*_0_.
- Scenario X.b: Increase *κ* = 5 of the baseline value *ω*_0_.
- Scenario X.c: Increase *κ* = 7 of the baseline value *ω*_0_.

where *X* = *I, II, III*. Fixed parameter values are given in Table 1. The length of the interval during which the compliance-failure rate is increased is 11 days (equation 2.4).

**Table 1:** Baseline parameter values for the simulation of the different scenarios described in the text.

| Parameter | Value | Parameter | Value | Parameter | Value |
| --- | --- | --- | --- | --- | --- |
| $b$ | 0.887677 | $\gamma^{-1}$ | 5.1 | $T$ | 30 |
| $\rho$ | 0.698725 | $\eta_a^{-1}$ | 6 | $q_{11}$ | 0.6 |
| $\mu$ | 0.11 | $\eta_s^{-1}$ | 7 | $\theta_1$ | 18 |
| $\omega$ | 0.005 | $\delta_s^{-1}$ | 4 | $q_{12}$ | 0.7 |
| $q$ | 0.7 | $\delta_r^{-1}$ | 12.070691 | $T^*$ | 67 |

### 2.1 (I) Long plateau with slight decrease and rebound

Setting *q*_1*i*_ as in Table 1, in (2.3) we put for both groups, *θ*_1_ of approximately two weeks and *θ*_2_ of about 3 months with *q*_21_ = 0.3 (decrease of 70% for the confined group), *q*_22_ = 0.4 (decrease of 60% for the unconfined group).

Figure 2 illustrates the case where the perturbed epidemic curve shows a slight decrease immediately after the perturbation relative to the baseline curve. Here, it is observed that as *κ* increases, the incidence increases again. Scenario *Ic* is the worst-case scenario where the growth rate keeps growing. Scenario *Ia* illustrates the appearance of a plateau-like behavior followed first by a slight decline and then growth again. The length of plateau is preserved for several weeks.

**Figure 2:**
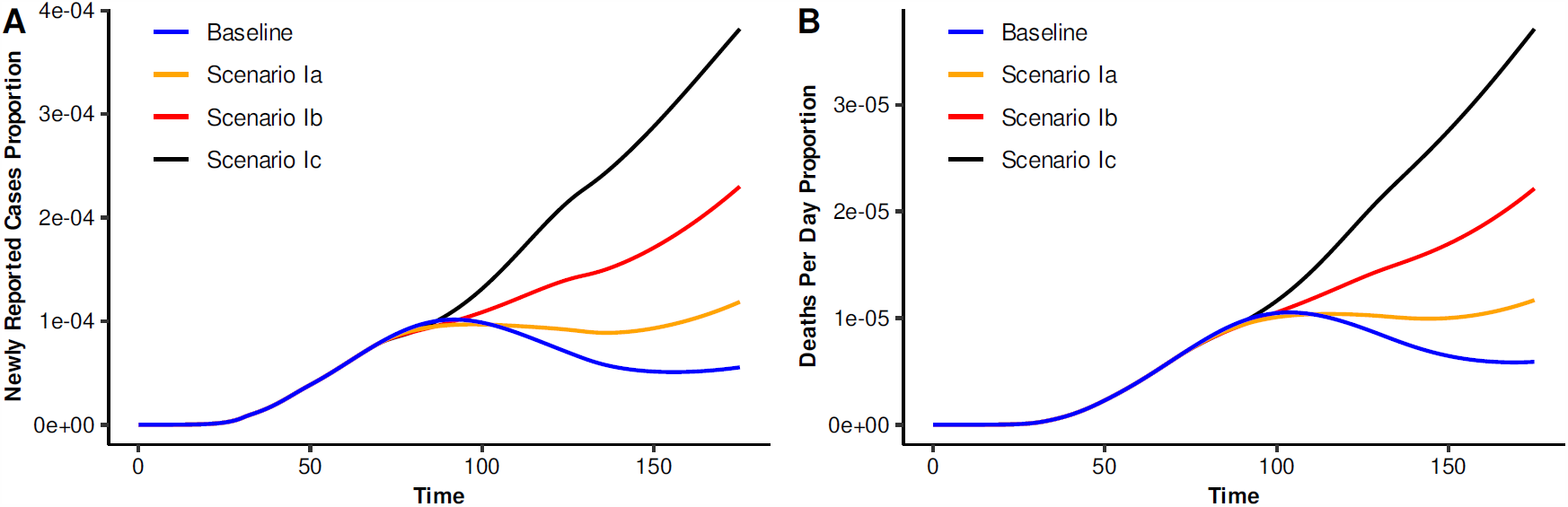
Impact of high mobility, for 12 days, on the baseline epidemic curve (blue line). The increase period starts 26 days before of the baseline epidemic curve peak. (A) Newly reported cases proportion per day, and (B) daily deaths proportion per day.

### 2.2 (II) Shorter plateau with decreasing epidemic-curve

Setting *q*_1*i*_ as in Table 1, in (2.3) we put, as before, *θ*_1_ of about two weeks and *θ*_2_ of about three months with *q*_21_ = 0.3 (decrease of 70% for the confined group), *q*_22_ = 0.3 (decrease of 70% for the unconfined group).

Figure 3 illustrates the case where the epidemic curve decreases after the end of the interval of maximum incidence. Scenario *IIc* is the worst-scenario since where after the perturbation, the epidemic peak is pushed higher and later in time. Scenario *IIa*, on the other hand, is comparatively benign. The peak is still reached as projected on the baseline case, and then the epidemic curve decreases at a slower rate than the baseline. Finally, scenario *IIb* shows the appearance of a plateau-like behavior. In this case, when the peak is reached, the incidence curve does not show a significant decay but rather, enters a sustained phase of maximum incidence that lasts several weeks.

**Figure 3:**
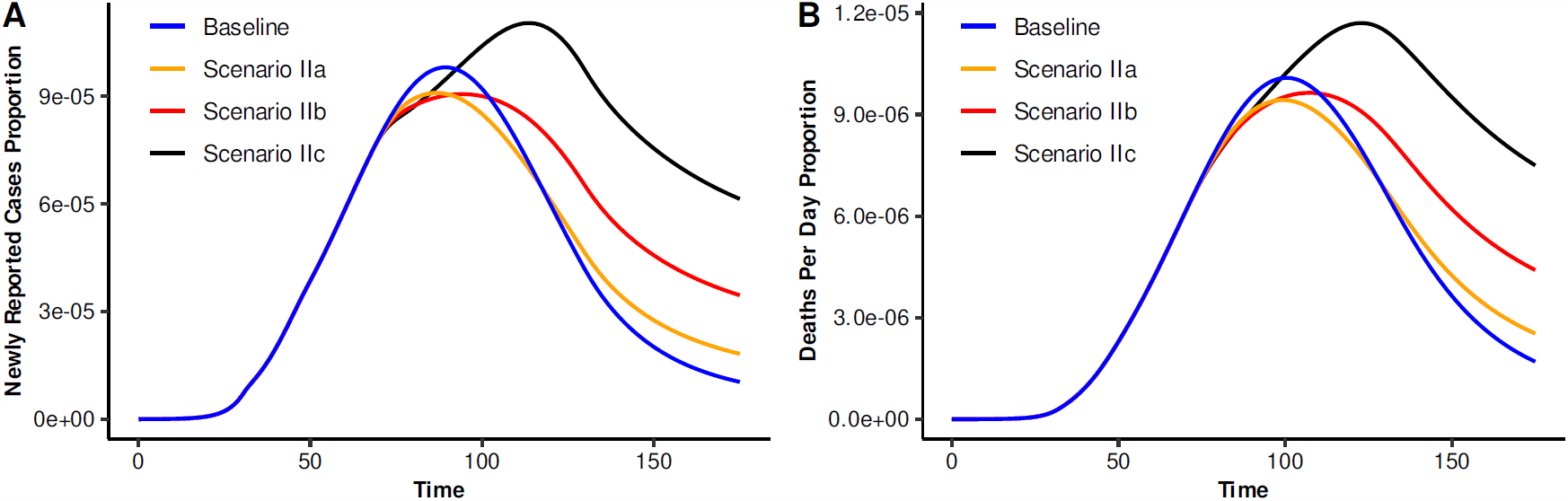
Impact of high mobility, for 12 days, on the baseline epidemic curve (blue line). The increase period starts 23 days before of the baseline epidemic curve peak. (A) Newly reported cases proportion per day, and (B) daily deaths proportion per day.

### 2.3 (III) Long plateau with decreasing epidemic-curve

Set *q*_1*i*_ as in Table 1, and, in (2.3) we put, for both groups, *θ*_1_ of about two weeks but now *θ*_2_ of about four months with *q*_21_ = 0.2 (confined group), *q*_22_ = 0.3 (non-confined group).

Figure 4 illustrate the case where the epidemic curve decreases after the peak. Scenario *IIIa* shows the appearance of a plateau that lasts some months after the date of the baseline peak. On the other hand, scenarios *IIIb* and *IIIc* shows a runaway epidemic with a much higher peak than that of the baseline curve.

**Figure 4:**
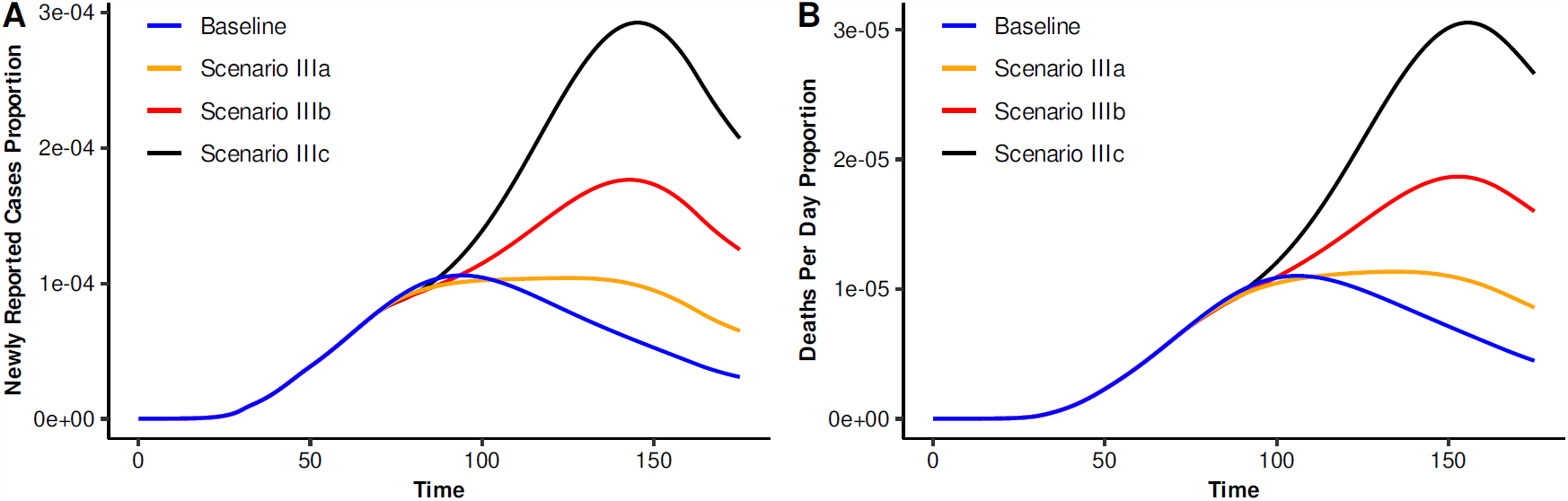
Impact of high mobility, for 12 days, on the baseline epidemic curve (blue line). The increase period starts 28 days before of the baseline epidemic curve peak. (A) Newly reported cases proportion per day, and (B) daily deaths proportion per day.

**Remark**. Depending upon the scenario, parameters *θ* _*j*_ and *q*_*i j*_ can be modified to explore alternative descriptions of it including the effective transmission contact rates (equation 2.3).

## 3 Case-study: Mexico City

In this section, we exemplify our model results with the case study of Mexico City. We start the analysis with some context. On March 23, 2020, social distancing measures where implemented in Mexico to slowdown the spread of COVID-19 pandemic, mainly focused in closing schools and some non-essential activities. One week later, on March 30, 2020, a Sanitary Emergency was declared to last until April 30, 2020, a date that was later extended to June 01, 2020. These measures aimed to “flatten the curve”, meaning to lower the incidence to ensure that critical cases would remain under manageable levels. On April 16, 2020, the federal government announced that for Mexico City, the day of maximum incidence would occur on May 08-10, 2020 [8, 9], and consequently, the date for lifting mitigation measures was announced to start on June 01, 2020. However, the peak did not happen on the projected dates. Independent projections by Mexican scientists [5, 6] set the dates of maximum incidence to be in an interval of about ten days around May 30, 2020, which puts the lifting of mobility restrictions indicated by the Federal government precisely in the likely days of higher transmission. Confronted with this scenario, the local Mexico City government puts into place a set of specific actions, that have kept the epidemic in the city within still manageable levels. Data shows that by the end of May, the epidemic in Mexico City seemed to stabilize on a plateau which continued beyond June up to the present day (mid July). This plateau may have appeared as a consequence of two important holidays, children’s day, and mother’s day, occurring approximately 10-14 days before the expected date for the peak of maximum incidence. We postulate that these events are an important factor that explains the observed quasi-stationary epidemic trend that the data shows at present (mid-July 2020). In this section we attempt an explanation of the probable causes of such behavior. Furthermore, we explore the possible consequences of lifting the mitigation measures under diverse conditions. COVID-19 epidemic data was provided by the Secretaría de Ciencia, Tecnología e Innovación of the Government of Mexico City through the COVID-19-CDMX database [10]. This data set contains details on all the confirmed and suspected COVID-19 individuals such as sex, age, residence place, date of symptoms onset, etc. We use records from February 22, 2020, to July 19, 2020.

### 3.1 Effect of mitigation measures

Richards model is an extension of a simple logistic growth model that is an standard tool commonly used to predict cumulative COVID-19 cases in China (see, for example, [11]). In this model the curve of cumulative cases, *C*(*t*), is described by the solution of

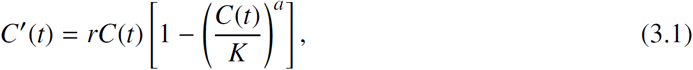

where *r* is the epidemic growth rate, *K* is the final epidemic size, and *a* is a parameter that accounts for the asymmetry of the epidemic curve. We estimate the Richards curve for two different periods, the first goes from February 22, 2020 to March 22, 2020, and the second from March 23, 2020 to April 30, 2020. The parameters *a, r*, and *K* for each period are estimated using Bayesian inference [12, 13]. Technical details can be found in Appendix A. We limit the fitting of the Richards curve to April 30 since on that date an important event of increased transmission started that violated the hypothesis that population conditions for transmission were essentially unchanged since March 23, 2020. In terms of our model, unchanged conditions mean *ω*(*t*) = *ω*_0_ for all *t*.

Before the start of social distancing on March 23, 2020, the growth rate is approximately 0.76 with a 95% interval (0.28, 1.91) which indicates a very fast growing epidemic with a doubling time of about a day. After March 23, 2020, the median growth rate is 0.12 with a 95% interval of (0.101, 0.168) rendering a median doubling time of about 6 days. This gives us a median reduction of 85% of the epidemic growth rate in the early days of the implementation of social distancing measures. The uncertainty around this value is significant but our estimate gives evidence that the confinement strategy was indeed effective in reducing transmission during the first weeks of mitigation. This justifies the idea of including decreasing effective contact rates. Unfortunately, however, those same social distancing measures also pushed the interval of the likely occurrence of the epidemic peak towards the end of May, thus overlapping it with the date set to reopen the economy.

**Remark:** The interval of dates for maximum incidence are in agreement in three previous models, for late May to early June. Two of them are mechanistic [5, 6]; the third is the projection of the Richards curve shown in Figure 5.

**Figure 5:**
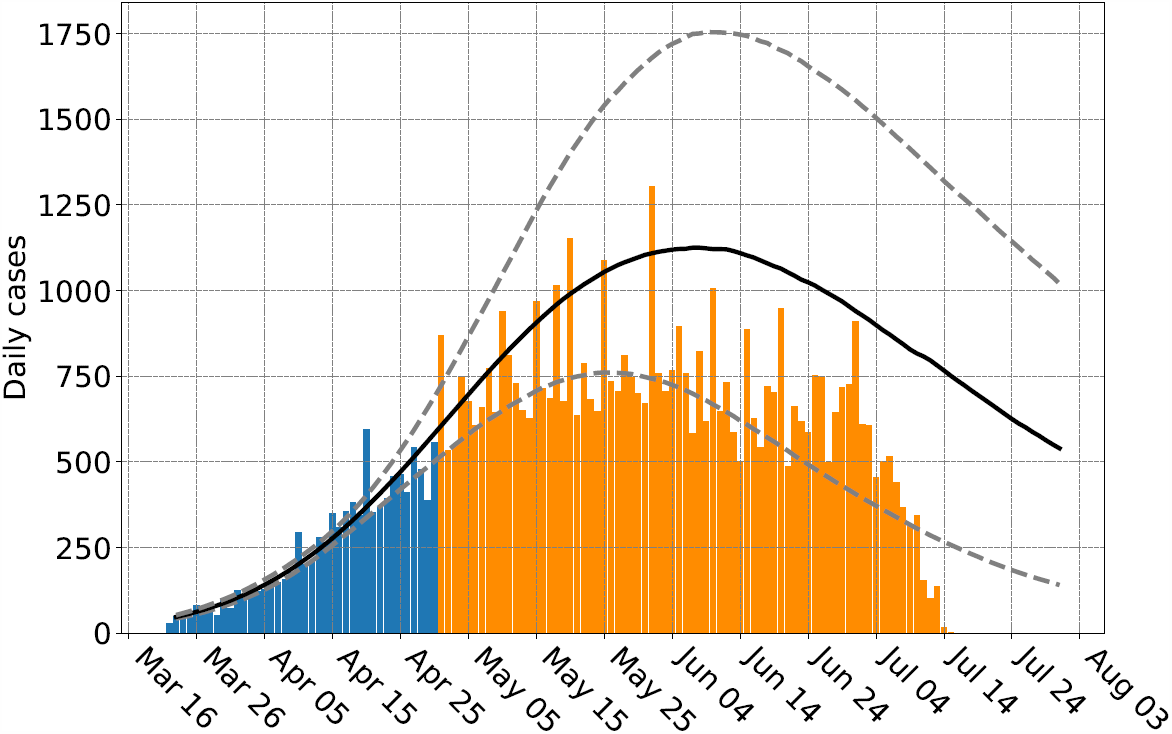
Number of daily new COVID-19 confirmed cases by symptoms onset and Richards model fit. The blue bars show data from March 23, 2020 to April 30, 2020 used to estimate the parameters. Orange bars show available data up to July 19, 2020 that were not used to estimate the parameters. Solid black line and gray dashed lines show pointwise median and 95% probability interval estimates for the expected number of cases, respectively. Note that the date of maximum incidence is projected to occur between May25 and June 8 (maxima of dotted lines). The projection is valid only up to May 30 since on June 01, confinement was partially lifted.

### 3.2 Mathematical model setup

To evaluate the impact of mobility events, we set-up equations 2.1-2.2 under the scenario that 70% of the general population is confined (obeying mitigation restrictions) during the confinement period (first time interval), and 30% is not [14]. The unconfined population includes workers with essential economic activities in government and industry and individuals that work in the informal economy. The time where confinement started *T* is March 23, 2020. The duration of the first reduction in the effective contact rate is *θ*_1_ = 18 days. The magnitudes of the first reduction in both effective contact rates are *q*_11_ = 0.6 (40% reduction in the confined group), *q*_12_ = 0.7 (30% reduction in the unconfined group), respectively. The duration *θ*_2_ and magnitudes (*q*_2*i*_) of the second reductions (*q*_2*i*_) depend on the study scenario as described in the next section.

Once under confinement, individuals may abandon the isolation with a compliance-failure rate *ω*(*t*). This is the parameter we use as a proxy for population mobility. *ω*(*t*) is a time-dependent rate: we assume that increased mobility lasts only for a period of *θ*_*ω*_ days, with a background compliance-failure rate *ω*_0_ that in atypical events is increased by a factor *κ* (equation 2.4). We take *ω*_0_ = 0.005*/*day as a baseline value [5], that is, given the exponential nature of this rate, this means that 50% of the confined populations will stay there at least for 4 months, while the other 50% will abandon confinement earlier.

## 4 Results

### 4.1 Increased mobility previous to the expected epidemic peak

Non-pharmaceutical interventions (NPIs) have been used to reduce contact between individuals in many countries. However, short-term increases in population mobility have been observed within the social-distancing confinement period. This increased mobility weakens the strength of the NPIs and, therefore, has an impact on disease transmission. Here we explore the consequences of increasing mobility during the confinement period. The objective is to give a plausible explanation for the appearance of the plateau in the dynamics observed in several countries using, as an example, the case of Mexico City.

In Mexico, there were two important holidays (in terms of population mobility) within the period of confinement: April 30, 2020, children’s day, and May 10, 2020, mother’s day. Population mobility increased these days as evidenced by the intantaneous reproduction number (Figure 6). Note that *R*_*t*_ shows a slight increase just around April 30, 2020, and May 01, 2020. We center our attention on the effect of increased mobility within the period from April 29, 2020, to May 10, 2020.

**Figure 6:**
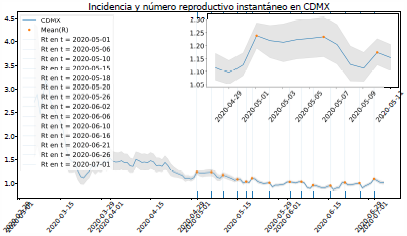
Instantaneous reproduction number for Mexico City using a median serial interval of 4.7 days [17]. The Figure shows the estimates from February 22, 2020 to July 7, 2020. In the insert a clear jump can be observed for April 30 and May 01, indicating a likely increase in active transmission during these days.

Mexico City epidemic has shown a plateau-like behavior since middle-May (Figure 8). As Figure 6 shows, a significant increase in mobility occurred in the period from April 29, 2020, to May 10, 2020. Therefore, we set the start of the perturbation, *T* ^∗^, as April 29, 2020, and its duration *θ*_*w*_ = 11 days. The second reductions in both effective contact rates are set to *q*_21_ = 0.3 and *q*_22_ = 0.4, while *θ*_2_ = 80 days. Other parameter values are fixed and given in Table 1.

Figure 7 shows simulations for *κ* = 3. We compare our results with the reported confirmed cases and deaths per day. Our projections go until July 01, 2020. Blue and yellow bars represent confirmed and suspect+confirmed cases, respectively. Observe that the mortality is not well described by our model (Figure 7). This observed decrease in mortality is intriguing. It could be due to enhanced treatment of grave cases, shifting of the morbidity towards age classes with a lower risk of death or, perhaps, incomplete mortality records and reporting time delays. Since we are comparing with the absolute number of deaths, testing has little impact in this case.

**Figure 7:**
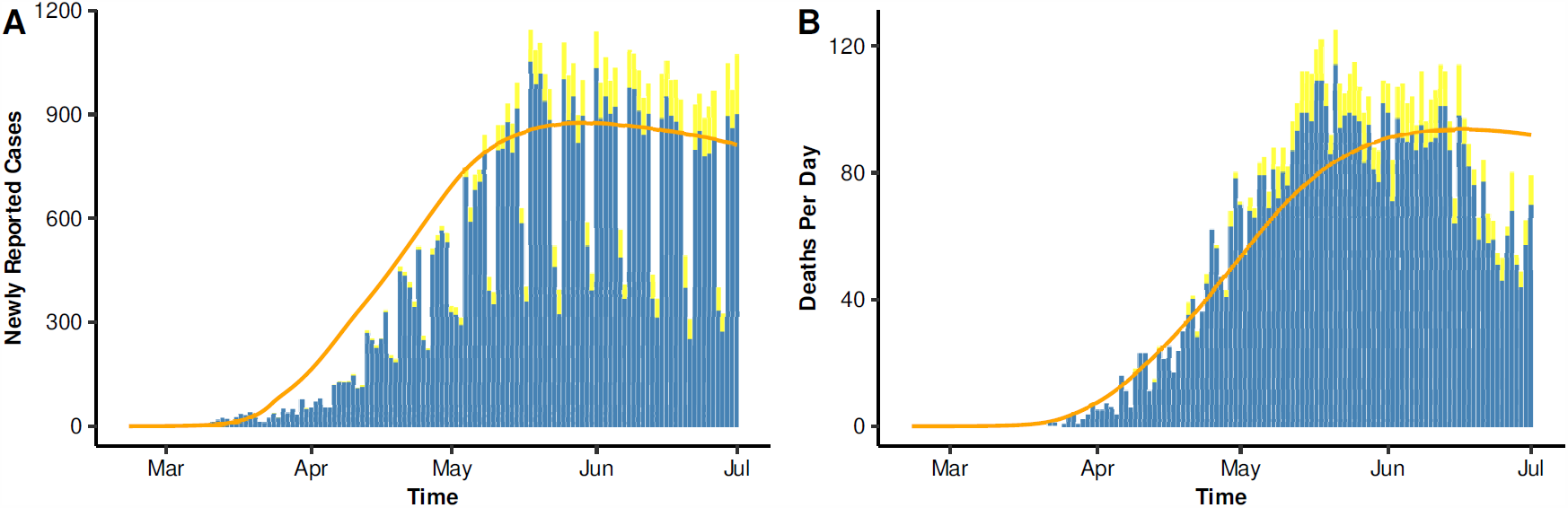
Impact of high mobility, from April 29, 2020, to May 10, 2020. (A) Newly reported cases per day, and (B) daily deaths per day.

### 4.2 Lifting social-distancing measures

Currently, an important question relates to the consequences of lifting too early the mitigation measures. The concern is that an increase in the number of cases may occur as it has occurred in the USA. In this section, we explore the possibility of this scenario for Mexico City.

Mexico’s federal government developed an epidemiological panel to oversee the reactivation of economic activities in the different states of the country. The panel is based on an index that essentially combines incidence and mortality data, and it has been used since June 01, 2020. The aim was to gradually increase the population mobility to keep under control the hospital demand and capacity. It has four colors: red, orange, yellow, and green.

At the time of writing (third week of July), the epidemiological panel for Mexico City has had two changes: it started under a red flag on June 01, 2020, the initial date for lifting of mobility restrictions; and it changed to an orange flag on June 29, 2020. A red flag allows only essential economic activities and brief trips to pharmacies, supermarkets, doctor, etc. An orange flag allows added non-essential activities with business staffed only to a 30% capacity, opening of public spaces with reduced capacity, etc [18, 19]. On both dates, an unknown number of individuals abandoned confinement and returned to their daily pre-epidemic activities.

We model this process by updating the number of individuals at a fixed time (*T* ^∗^) in equation (2.2), as *x*_1_(*t*_*i*_) = (1 − *q*)*x*_1_(*t*_*i*−1_) and *x*_2_(*t*_*i*_) = *qx*_1_(*t*_*i*−1_) + *x*_2_(*t*_*i*−1_) where *t*_*i*_ = *T* ^∗^, *x*_1_ and *x*_2_ represent the susceptible, exposed, asymptomatically infected and symptomatically infected for the confined and unconfined groups, respectively.

#### 4.2.1 Lifting mitigation measures on June 29, 2020

Now we explore the possible consequences of lifting mitigation measures on June 29, 2020. We have chosen this date based on an actual change of the Mexico City epidemiological panel (a change from red to orange light). We explore the short and medium-term effects of the lifting of the mitigation measures on the epidemic curve. For each case, we project three scenarios: 25%, 50%, or 75% of the confined population abandoning confinement. We present three curves that have a good fit to Mexico City data. These curves are obtained for the parameters shown in Table 2.

**Table 2:** Parameters for the lifting of mitigation measures scenarios.

| Parameter | Blue line | Red line | Black line |
| --- | --- | --- | --- |
| $b$ | 0.887677 | 0.906658 | 0.911977 |
| $\rho$ | 0.698725 | 0.644263 | 0.660068 |
| $\delta_r^{-1}$ | 12.070691 | 13.838454 | 12.263355 |

Both effective contact rates (for the confined and not-confined groups) decrease until June 29, 2020, and then both remain constant. For this scenario, reductions in effective contact rates are described by *q*_11_ = 0.6, *q*_21_ = 0.3 (for the confined group), and *q*_12_ = 0.7, *q*_22_ = 0.4 (for the not-confined group).

Figure 8 shows the behavior of reported cases *I*_*r*_. Solid lines are the baseline, while dotted lines represent the perturbed system (after lifting restrictions on June 29, 2020). Note that, as the percentage of the confined population is decreased, the incidence increases to higher levels. Also, for the scenario of 25% of the population leaving confinement (i.e., increasing its effective contact rate), we obtain a good data fit.

#### 4.2.2 Lifting on June 29, 2020, with decreasing effective contact rate

We explore the scenario where the effective contact rate continues to decline even after partially lifting mitigation measures. Here, we consider that the effective contact rates in both groups decrease until August 01, 2020, and then both remain constant. For this scenario, we set *q*_11_ = 0.6, *q*_21_ = 0.1 (confined group), and *q*_12_ = 0.7, *q*_22_ = 0.2 (unconfined group). Other parameter values are similar to those employed in the above subsection.

Figure 9 underlines the importance of a continuing decrease of the transmission rate after lifting. Although the contingency measures are lifted on June 29, in the worst-case scenario (Figure 9D), there can be a reduction of approximately 50% of newly reported cases (see Figure 8D) after lifting the mitigation measures.

**Figure 8:**
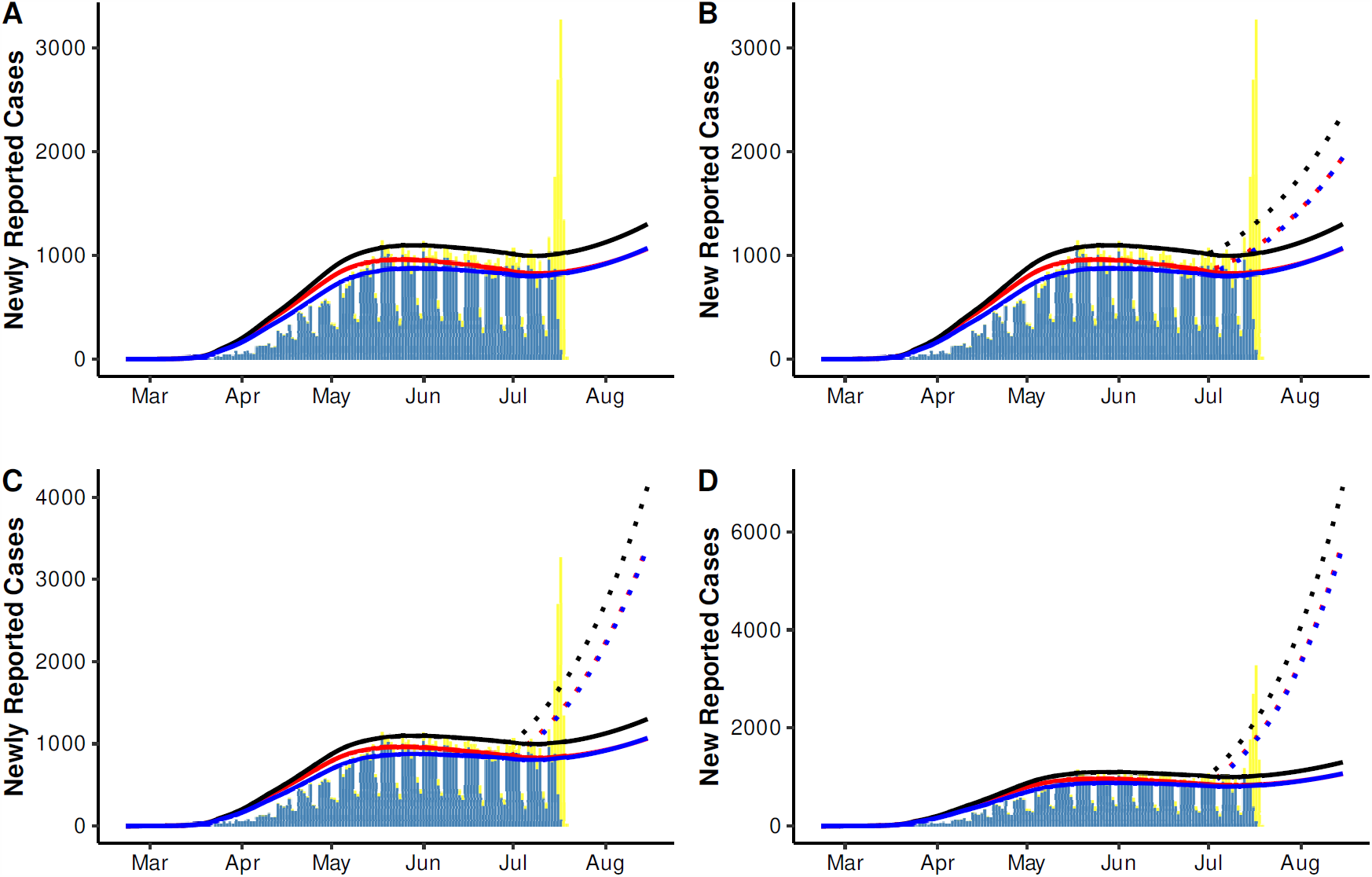
Lifting mitigation measures on June 29, 2020. A) Dynamics, when confinement is not lifted, B) dynamics when 25% of the confined population leaves confinement, C) when 50% of the confined population leaves confinement, and D) if 75% of the confined population leaves confinement.

**Figure 9:**
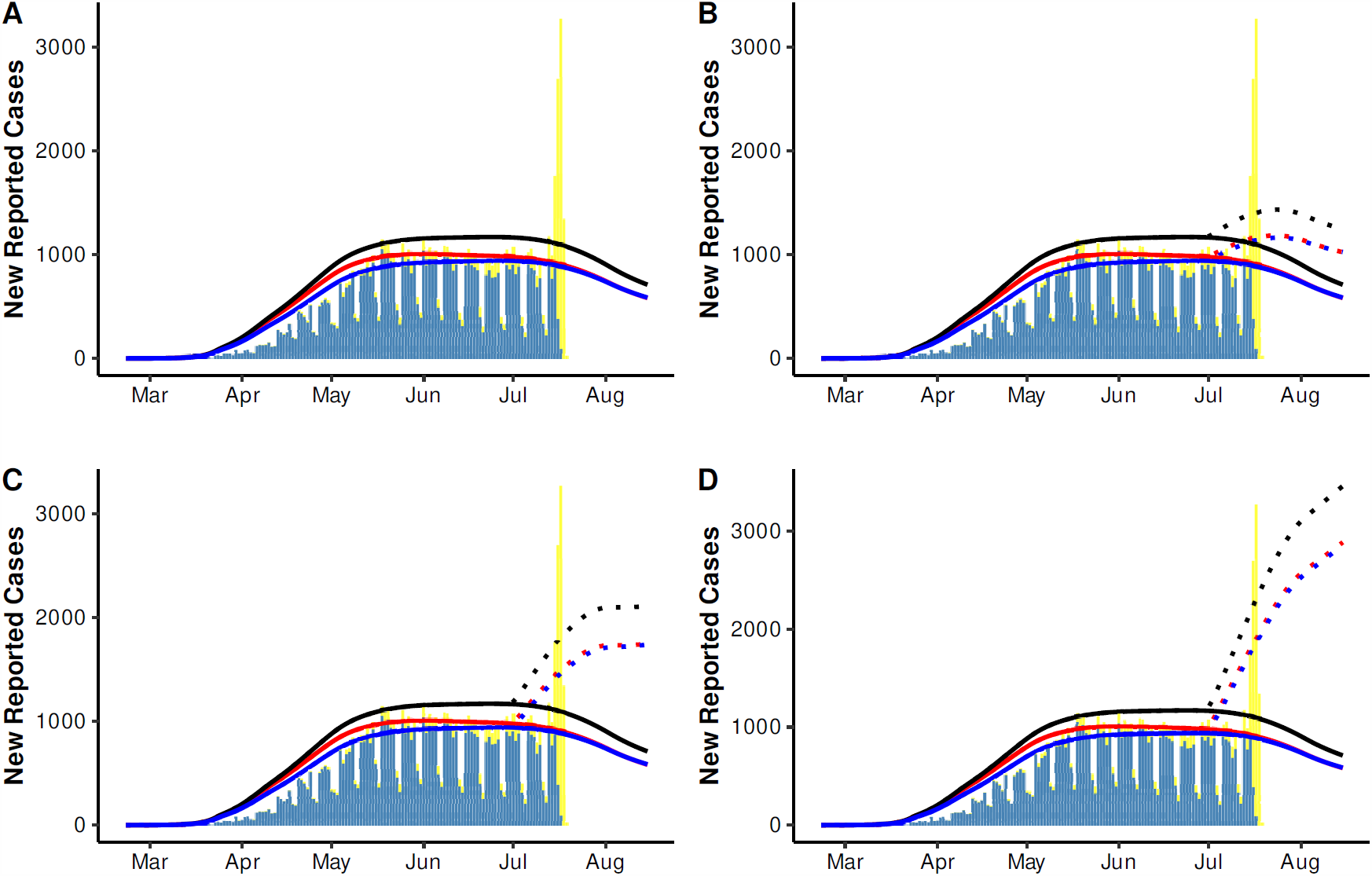
Lifting of the mitigation measures on June 29, 2020, but both effective contact rates (for lockdown and non-lockdown environments) decrease until August 01, 2020. A) Dynamics, when nobody leaves confinement, B) dynamics when 25% of the confined population leaves confinement, C) if 50% of the confined population leaves confinement, and D) when 75% of the confined population leaves confinement.

##### Remark

Figures 8 and 9 show that, as the percentage of the population under confinement decreases, the number of new reported cases also increases. A similar behavior is observed for the dynamics of daily deaths.

## 5 Conclusion

The analysis of the precise epidemiological situation in Mexico City is difficult because, among other things, COVID-19 testing is limited, the positivity rate is high and there is substantial underreporting of cases and mortality. Our case-study uses the confirmed cases reported by the Mexico City government in its official web page. Results show that the growth rate of the epidemic was reduced upon the implementation of mitigation measures on March 23, 2020. However, for Mexico City, the federal government originally forecasted the peak of maximum incidence to occur by May 8-10, 2020 and the date for lifting mitigation measures was set to start on June 1, 2020. The peak, however, occurred much later in the month as the data and independent models have shown [5, 6, 7]. The epidemic in Mexico City shows that it entered a period of constant incidence since around May 20 that, as mentioned in the previous sections, still continues. The end result was that the lifting of mobility restrictions coincided with this interval of maximum incidence. Events of high mobility and increased transmission occurring during confinement on times relatively close but before the expected epidemic peak (e.g., like children’s day and mother’s day in Mexico City), may have impacted the epidemic curves pushing it into the plateau behavior observed in the data and described in the different scenarios presented in this work. We have explored the effect of pulses of unusual activity (within the confinement period) and their effect of the lifting on the epidemic curve of COVID-19 in Mexico City. These are theoretical results, but we believe, they illuminate the importance of counting with reasonable estimates for the timing of maximum incidence and of constant surveillance and evaluation of atypical high transmission events during confinement. A characteristic of our model that we think is worth underlining, is that the effective contact rates are time-varying which, we think, constitutes a realistic approximation since we are looking at population averages. For example, adopting safe behaviors, such as wearing face masks, involves a learning process; it does not suddenly occur, rather it takes time for the face masks to be adopted by a significant proportion of the general population. Our results indicate that after lifting mobility restrictions, a decrease of the effective contact rate should continue in order to force the epidemic curve to make a downward turn. We interpret this continuing decrease in the effective contact rate as related to the use of face masks, social-distancing, washing hands, etc.

Mathematical models are essential in the fight against COVID-19. They are tools for evaluating mitigation measures, estimating mortality and incidence, and projecting scenarios to help public health decision-makers in their very difficult and important task of controlling the epidemic. In this paper, we have used mathematical models to evaluate and generate scenarios. Although precise forecasting is not our aim, we consider that these results can be helpful for decision-makers.

## Data Availability

We declare that COVID-19 data used in this paper is available online.

https://www.gob.mx/salud/documentos/datos-abiertos-152127

## Acknowledgments

All authors acknowledge support from DGAPA-PAPIIT-UNAM grant IV100220 (convocatoria especial COVID-19).JXVH acknowledges support from grant PAPIIT-DGAP IN115720 We thank Dr. Héctor Benitez, and Dr. William Lee Ardavín for their support. We also thank the Secretaría de Ciencia, Tecnología e Innovación of the Government of Mexico City for facilitating the data on the COVID-19 epidemic.

## A Appendix A: Richards curve estimation

Let *Y*_*j*_, for *j* = 1, 2, …*n*, be the number of observed new daily cases at time *t* _*j*_, with *t* _*j*_ given in days since the first reported case started symptoms. We assume that *Y*_*j*_ follows a Negative Binomial distribution with mean value *X*(*t*_*j*_|*a, r, K*) = *C*(*t*_*j*_|*a, r, K*) − *C*(*t*_*j*−1_|*a, r, K*) and dispersion parameter *α*. Here, *C*(*t*|*a, r, K*) is the solution of Richards model presented in (3.1). Assuming that, given the parameters, the observations *Y*_1_, *Y*_2_, …, *Y*_*n*_ are conditionally independent, then

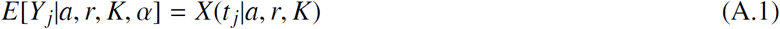

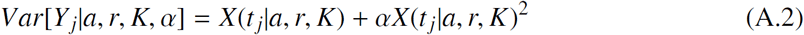

The Negative Binomial distribution allows to control the variability of the data by considering over-dispersion which is common for epidemiological data. If *α* = 0, then we return to the Poisson model which is often used in this context.

Let θ = (*a, r, K, α*) be the vector of parameters to estimate. The inclusion of the parameter *α*, which is related to the variability of the data, not to the Richards model, is necessary since in practice this variability is unknown. Then, the likelihood function, which represents how plausible is the data under the Negative Binomial assumption and Richards model if we knew the parameters, is given by

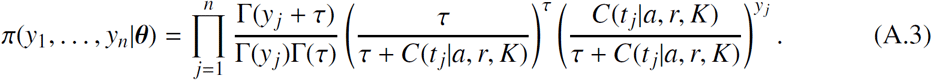

Consider that parameters *a, r, K* and *α* as random variables. Assuming prior independence, the joint prior distribution for vector θ is

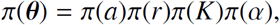

where *π*(*a*) is the probability density function (pdf) of a Uniform(0,2) distribution, *π*(*r*) is the pdf of a Uniform(0, 2), *π*(*K*) is the pdf of a Uniform(*K*_min_, *K*_max_), and *π*(*α*) is the pdf of a Gamma(shape=2, scale=0.1). To select the prior for parameter *r*, we consider that previous estimations of *r* are close to 0.3 [11], and a Uniform(0,2) represents a weekly informative prior as it allows for a wide range of values of *r*. Also, there is no available prior information regarding the final size of the outbreak *K*. This is a critical parameter in the model and, in order to avoid bias, we assume a uniform prior over *K*_min_ and *K*_max_. To set these last two values, we consider that the minimum number of confirmed cases is the maximum number of observed cumulative cases *Y*(*t*_*n*_) (for the studied period) times two, i.e. *K*_min_ = *y*_*n*_ ∗ 2. To set the upper bound for *K*, we consider a fraction of the total population *K*_max_ = *N* ∗ 0.05, where *N* is the population size of Mexico. This fraction was determined based on the observations of other cities such as the New York where the total population size is similar to Mexico City.

Then, the posterior distribution of the parameters of interest is

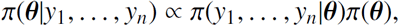

and it does not have a simple form since the likelihood function depends on the solution of the Richards model, which is not linear on the parameters. We analyze the posterior distribution using an MCMC algorithm that does not require tuning called *t-walk* [20]. This algorithm generates samples from the posterior distribution that can be used to estimate marginal posterior densities, mean, variance, quantiles, etc. We refer the reader to [12] for more details on MCMC methods and to [13] for an introduction to Bayesian inference with differential equations.

We estimate vector θ using data from Mexico City for two different periods, from February 22, 2020 to March 22, 2020, and from March 23, 2020 to April 30, 2020. Table 3 shows the median posterior estimates and 95% probability intervals for the parameters in each period.

**Table 3:** Parameter median estimates and 95% posterior probability intervals for Richards model. Two periods are considered, from February 22, 2020 to March 22, 2020, and for two different periods, from March 23, 2020 to April 30, 2020. Here, *r* is the growth rate, *K* is the final size of the outbreak and *a* is a scaling factor. *K* is shown by completeness.

|  | February 22 - March 22 |  |  | March 23 - April 30 |  |  |
| --- | --- | --- | --- | --- | --- | --- |
|  | Lower | Median | Upper | Lower | Median | Upper |
| $a$ | 0.02 | 0.05 | 0.38 | 0.12 | 0.17 | 0.24 |
| $K$ | 1,186 | 26,156 | 167,504 | 31,737 | 67,344 | 88,417 |
| $r$ | 0.28 | 0.76 | 1.91 | 0.10 | 0.15 | 0.26 |

